# Thalamic, hippocampal, amygdalar and brainstem long-term changes in schizophrenia co-localised with normative neurotransmitter distributions

**DOI:** 10.64898/2026.09.21.26363537

**Authors:** Claudio Alemán-Morillo, Natalia García-San-Martín, Richard AI Bethlehem, Patricia Segura, Chloe Gomez, Alessia Pasquini, Pablo Salguero-Quirós, Manuel Muñoz-Caracuel, Rosa Ayesa-Arriola, Javier Vázquez-Bourgon, John Suckling, Miguel Ruiz-Veguilla, Benedicto Crespo-Facorro, Rafael Romero-García

**Author notes:** Correspondence to: Rafael Romero García, Avda. Doctor Fedriani S/N, 41009, Seville (Spain).

## Abstract

**Background:** Schizophrenia Spectrum Disorders (SSD) is associated with spatially heterogeneous structural abnormalities across distributed brain systems, but the longitudinal evolution and neurochemical context of subcortical subregional changes remain poorly understood. We investigated longitudinal volume changes across thalamic, hippocampal, amygdalar, and brainstem subregions and their relationships with clinical characteristics and normative neurotransmitter distribution.

**Methods:** A total of 357 SSD individuals with first-episode and 196 healthy controls were included. Clinical assessments were performed at baseline and at 1, 3, 5, 10, 15, and 20 years of follow-up. Using T1-weighted structural MRI, subregional volumes were quantified across the thalamus, hippocampus, amygdala, and brainstem. We examined baseline group differences, longitudinal volume changes, and associations with clinical variables, including symptom severity. Spatial patterns of regional associations were compared with normative maps of neurotransmitter receptor and transporter distributions.

**Results:** Patients showed heterogeneous structural alterations across the examined subcortical subregions. The hippocampus showed prominent early volume deficits in specific subfields and demonstrated a strong association with antipsychotic medication doses. Amygdalar volumetric reductions were more widespread and were strongly associated with positive symptom severity. Localised structural abnormalities were also observed in the thalamus and brainstem, although these showed weak associations with clinical symptomatology. The spatial co-location of normative neurotransmitter maps with the structural and clinical associations revealed distinct overlapping with dopaminergic, GABAergic, glutamatergic, serotonergic and cholinergic systems.

**Conclusions:** Longitudinal structural alterations in SSD showed marked anatomical heterogeneity across thalamic, limbic, and brainstem systems. Their spatial correspondence with normative neurochemical architecture suggests that regional and subregional structural vulnerability and clinical heterogeneity may be organized along distinct, partially overlapping neurotransmitter profiles.

## INTRODUCTION

Current neurobiological models of schizophrenia increasingly emphasize dysfunction across distributed brain circuits rather than isolated regional abnormalities [1]. Within these networks, interconnected thalamic [2–4], limbic [5, 6], and brainstem [7–10] systems are of particular interest because of their involvement in areas as cognition and emotional processing and their modulation by neurotransmitter systems critically implicated in the pathophysiology of schizophrenia [11–17]. Growing evidence suggests that abnormalities within these systems are spatially heterogeneous, preferentially affecting specific subregions with distinct connectivity profiles and neurochemical properties [11, 12, 18]. This regional heterogeneity highlights the importance of examining subcortical structures at the subregional level, rather than treating them as anatomically and functionally homogeneous units.

The thalamus is a major integrative hub linking cortical and subcortical networks [2]. Studies in schizophrenia have frequently reported alterations in thalamocortical connectivity, including reduced connectivity with prefrontal regions and increased connectivity with sensorimotor cortices [19]. Structural abnormalities also appear to preferentially involve specific thalamic nuclei, particularly the mediodorsal nucleus [3, 20], which is strongly connected with the prefrontal cortex [21], and the pulvinar [22–24], although alterations in lateral and ventral nuclei have also been found [25, 26]. Volumes of the mediodorsal nucleus and pulvinar have been associated with cognitive performance in schizophrenia [20, 24], whereas auditory hallucinations have been related to structural alterations across several thalamic territories, including the medial, lateral geniculate, pulvinar, and mediodorsal nuclei [27].

The hippocampus is similarly implicated in the pathophysiology of schizophrenia, with converging evidence of structural and functional abnormalities [28–31]. Hippocampal volume has been associated with negative symptoms [29], whereas alterations in specific subfields, including CA2/3 and CA4/dentate gyrus, have been related to deficits in visual and working memory [32]. Beyond these clinical and cognitive associations, the hippocampal hyperactivity model [33] provides a potential neurochemical framework for understanding hippocampal dysfunction in schizophrenia. The model proposes that GABAergic interneuron dysfunction leads to impaired inhibitory control and subsequent pyramidal-cell disinhibition, resulting in hippocampal hyperactivity. This hyperactivity is proposed to increase hippocampal output and, in turn, dysregulate mesolimbic dopaminergic signaling, providing a potential mechanistic link between hippocampal dysfunction and psychotic symptoms.

The amygdala represents another key component of these interconnected systems because of its central role in emotional processing, particularly in fear- and anxiety-related processes [34]. Structural [35, 36] and functional [37] abnormalities of the amygdala have been associated with psychotic symptoms and impaired emotion recognition [37]. As in the thalamus and hippocampus, these alterations may show subregional specificity. For example, auditory hallucinations have been associated with alterations in the left accessory basal nucleus [38], whereas centromedial amygdala volume has been related to anhedonia [39]. At a circuit level, the basolateral amygdala has been proposed to contribute to schizophrenia pathophysiology through abnormal modulation of interconnected corticolimbic regions, potentially disrupting GABAergic inhibitory regulation in the hippocampus and anterior cingulate cortex [40, 41].

Although brainstem morphology has been less extensively studied, it is also a major contributor to the pathophysiology of schizophrenia. Midbrain dopaminergic nuclei receive inputs from the hippocampus, thalamus, and prefrontal cortex and provide widespread dopaminergic modulation of striatal and cortical regions. Dysregulation of these circuits has been proposed to underlie the abnormal dopaminergic signaling associated with psychosis [8]. Beyond its role in dopaminergic signalling, the brainstem comprises the principal serotonergic, noradrenergic, and cholinergic nuclei, implicating multiple neuromodulatory systems in schizophrenia [42].

Together, this evidence indicates that structural abnormalities in schizophrenia are regionally heterogeneous and may reflect the brain’s underlying neurochemical organization. However, it remains unclear whether this regional heterogeneity extends to the temporal domain, with different subregions showing distinct trajectories of volumetric change over the course of illness, and whether these longitudinal patterns are associated with the normative molecular architecture of the brain. Furthermore, the extent to which longitudinal subregional changes track changes in clinical symptom severity remains poorly understood. Addressing these questions may provide insight into the molecular organization and clinical relevance of longitudinal subcortical alterations in schizophrenia.

In the present longitudinal study, we investigated patterns of volume loss across subregions of the thalamus, hippocampus, amygdala, and brainstem, and examined their associations with longitudinal changes in clinical symptoms. We further tested whether the spatial distribution of longitudinal volume loss was co-located with normative maps of neurotransmitter receptor and transporter distributions. We hypothesized that these structural alterations would exhibit a non-random spatial organization consistent with regional molecular architecture and would be associated with concurrent changes in clinical symptom severity.

## MATERIAL AND METHODS

### Subjects

Participants were recruited through the Program for Attention to the Initial Phases of Psychoses (PAFIP), an intervention service established at Marqués de Valdecilla University Hospital in 2001. The study included individuals experiencing a first episode within the schizophrenia spectrum and neurotypical control participants. At study entry, most patients had not received antipsychotic treatment, although a subset of 40 participants received minimal antipsychotic medication prior to enrolment, with a mean treatment duration of 9±9.72 days (range 0-35 days).

The sample consisted of 195 healthy controls (120 males; mean age = 29.1 ± 7.63 years) and 357 patients with SSD (213 males; mean age = 29.8 ± 8.76 years, SAPS score =13.3 ± 5.27, SANS score = 5.89 ± 5.70). All patients fulfilled DSM-IV criteria for one of the diagnoses included under the SSD umbrella. The distribution of patients across diagnoses was as follows: schizophrenia (n = 213), schizophreniform disorder (n = 47), brief psychotic disorder (n = 37), psychotic disorder not otherwise specified (n = 24), schizoaffective disorder (n = 34), or delusional disorder (n = 2). Diagnostic status was reassessed after a 6-month follow-up period to confirm the initial diagnosis. Although the SSD group included several diagnoses, these were analyzed collectively based on evidence of shared neurobiological and psychopathological features across disorders [43–47]. Detailed inclusion and exclusion criteria are provided in the Supplementary Methods. Demographic and clinical characteristics of the sample, stratified by sex and diagnosis, are reported in Tables S1, S2 and S3. of the Supplementary Material.

The study was conducted in accordance with the Declaration of Helsinki and received approval from the CEIC-Cantabria Ethics Committee (clinical trial numbers NCT0235832 and NCT02534363), the University of Seville Ethics Committee (SICEIA 2024-2534) and the Hospital Universitario Virgen Macarena-Virgen del Rocío (SICEIA-2024-002682). Written informed consent was obtained from all participants prior to study enrollment.

### MRI T1 acquisition

Structural MRI data were acquired using two scanners: a 1.5T General Electric SIGMA system (GE Healthcare, Milwaukee, WI, USA) and a 3T Philips Achieva scanner (Philips Medical Systems, Best, The Netherlands) equipped with an 8-channel head coil. To ensure consistency across longitudinal assessments, all scans from a given participant were acquired on the same scanner.

For the 1.5T GE system, T1-weighted images were acquired with repetition time (TR) = 24 ms, echo time (TE) = 5 ms, flip angle = 45°, number of excitations (NEX) = 2, field of view (FOV) = 26 × 19.5 cm², matrix size = 256 × 192, slice thickness = 1.5 mm, and voxel dimensions = 1.02 × 1.02 × 1.5 mm³. For the 3T Philips system, acquisition parameters were TR = 8.2 ms, TE = 3.7 ms, flip angle = 8°, matrix size = 256 × 256, voxel dimensions = 0.94 × 0.94 × 1 mm³, and 160 contiguous slices.

Image quality was assessed visually. Scans were reviewed by trained raters and classified as pass, borderline, or fail according to the presence and severity of motion artefacts. Scans exhibiting substantial artefacts, including ghosting, blurring, or ringing, were classified as fail and excluded from further analyses. Mean Euler numbers were included as a quantitative proxy for motion-related artifacts among the retained scans (pass and borderline) and entered as a covariate in the statistical models.

Structural images were processed using longitudinal FreeSurfer [48] version 7.4.1. To account for scanner-related effects, volumetric measures were harmonized across scanners using ComBatLS [49] while preserving variance associated with age, sex, and diagnostic group.

### Subcortical volumetric segmentations

Subregional segmentation was performed using the longitudinal FreeSurfer subregion segmentation framework (https://surfer.nmr.mgh.harvard.edu/fswiki/SubregionSegmentation) for thalamic nuclei, hippocampal subfields, amygdalar nuclei, and brainstem structures (Figure 1). The segmentation procedures relied on probabilistic atlases and Bayesian inference methods for automated labeling of structural MRI data according to the longitudinal pipeline implemented in FreeSurfer, in which all timepoints are jointly incorporated through subject-specific atlas estimation [50]. The thalamic atlas was generated from manual delineations of 26 thalamic nuclei on serial histological sections from postmortem samples combined with in vivo MRI segmentations [51]. Hippocampal subfield segmentation was based on a computational atlas derived from ex-vivo MRI acquisitions of autopsy samples at ultra-high isotropic resolution with manual labeling [52]. Amygdalar nuclei segmentation relied on an ex-vivo probabilistic atlas generated from high-resolution 7T MRI scans with manual delineation [53]. Brainstem segmentation was performed using a probabilistic atlas generated from manually labeled MRI datasets [54].

**Figure 1.**
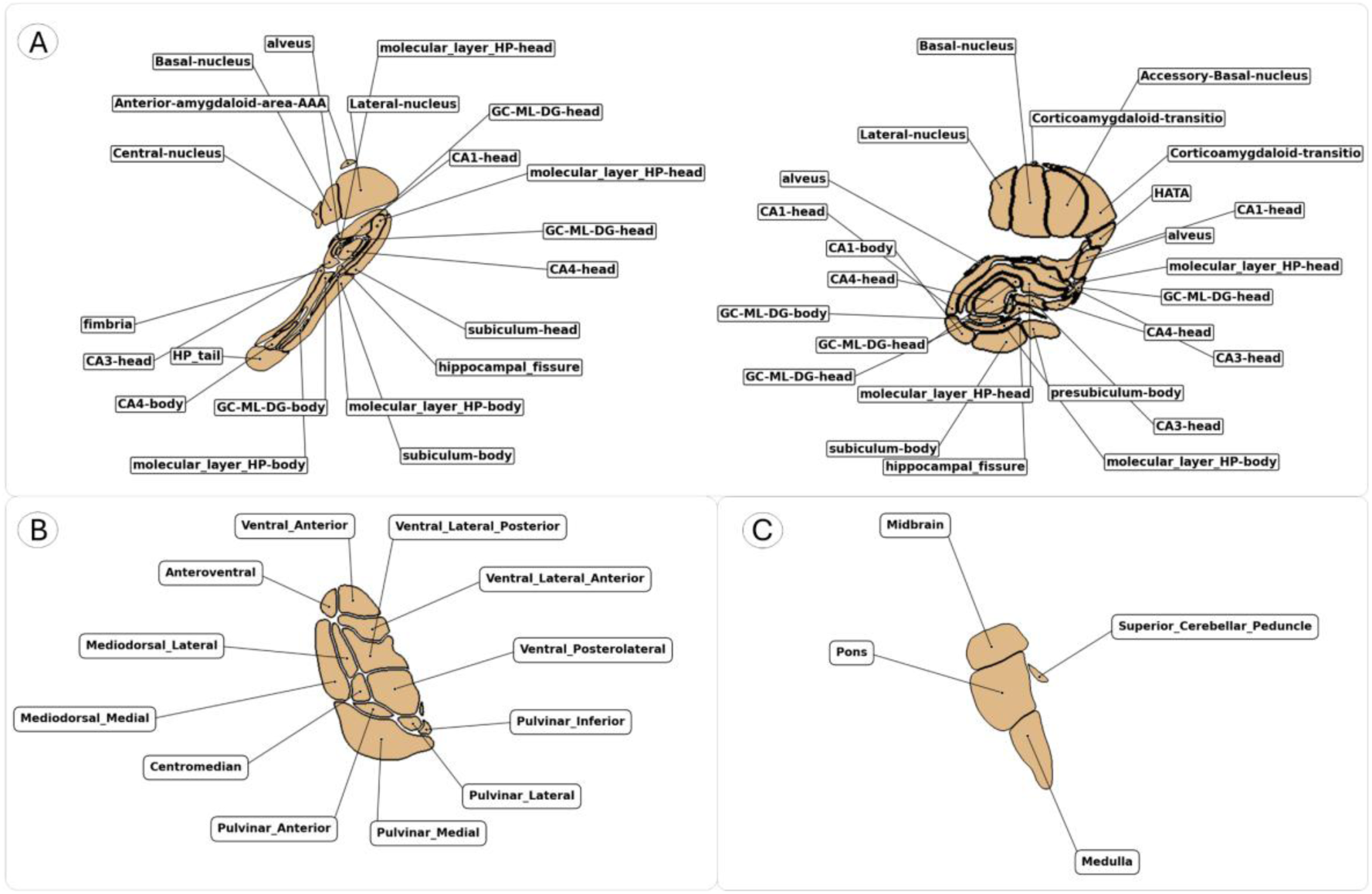
Subregional segmentation of the hippocampus, amygdala, thalamus and brainstem. Brain sections are displayed in MNI152 space: (A) hippocampus and amygdala (sagittal = ± 27 mm, axial = −17 mm), (B) thalamus (axial = 6 mm), and (C) brainstem (sagittal = −5 mm).

### Clinical assessments

Positive and negative symptoms in the SSD group were assessed with the Scale for the Assessment of Positive Symptoms (SAPS) and the Scale for the Assessment of Negative Symptoms (SANS) [55–57].

As this was a long-term prospective study, the potential impact of attrition bias was assessed by comparing baseline symptom severity between patients who remained in the study for at least 5 years of follow-up and those who dropped out before that time point. Attrition study and attrition data are available in supplementary material.

### Neurotransmitter distribution maps

A total of 38 neurotransmitter maps in MNI-152 space were obtained from NeuroMaps [58]. Specifically, the evaluated targets comprised acetylcholine receptors (α_4_β_2_ and muscarinic M_1_), the vesicular acetylcholine transporter (VAChT), cannabinoid receptor type 1 (CB_1_), dopamine receptors (D_1_ and D_2_), dopamine transporter (DAT), GABA receptor, metabotropic glutamate receptor 5 (mGluR_5_), histamine receptor H_3_, μ-opioid receptor (MOR), norepinephrine transporter (NET), serotonin receptors (5-HT_1A_, 5-HT_1B_, 5-HT_2A_, and 5-HT_6_), and the serotonin transporter (5-HTT). When multiple PET datasets were available for a given molecular target, each map was included as an independent observation. A complete list of maps used is shown in supplementary material (Table S4).

### Association between SSD diagnosis and subregional volumes

Cross-sectional associations were assessed using general linear models (GLMs), with the following model fitted separately for each brain region:

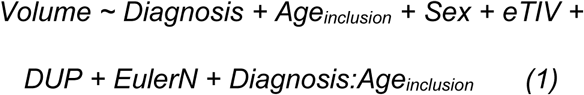

Where *Diagnosis* is a binary variable coded as 0 for control and 1 for cases, *Age_inclusion_* is the age at baseline MRI, *Sex* is included as a biological covariate (coded as 0 = male, 1 = female), *eTIV* (estimated total intracranial volume) to account for individual differences in head size, *DUP* is the Duration of Untreated Psychosis, *EulerN* is the mean Euler number derived from the cortical surface reconstruction, included as a proxy for image quality. The interaction term (*Diagnosis:Age_inclusion_*) was included to assess whether the association between age at enrolment and regional brain volume differed between groups.

Longitudinal effects on regional volume were evaluated using linear mixed-effects models, with each subject included as a random effect to account for repeated measurements.

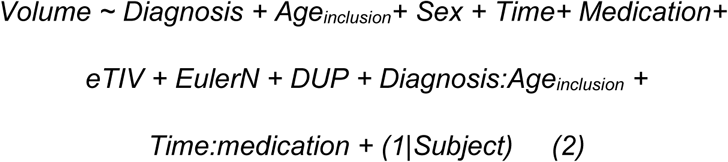

Where *Time* represents the period since the enrolment for controls, or since treatment initiation for participants with SSD, *Medication* is the dose of antipsychotic medication expressed in chlorpromazine equivalents (healthy controls were assigned a value of 0 for modelling purposes), *Time:Medication* captures whether medication alters the effect of treatment time in regional volumes.

All non-binary variables were *z*-scored, and statistical significance was assessed after correction for multiple comparisons using False Discovery Rate (FDR), with corrected p-values < 0.05 considered significant.

### Association between symptoms severity and subregional volumes

Linear mixed-effects models were fitted separately for total positive and negative symptoms, with each subregional brain volume entered as a predictor in region-specific models:

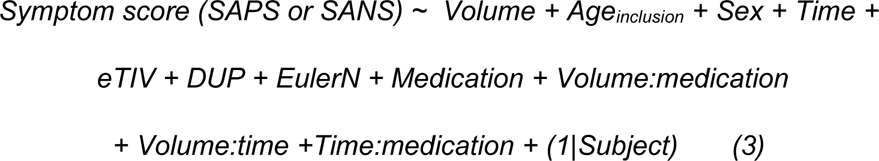

where the dependent variable was the symptom score, corresponding to either SAPS or SANS.*Volume* represents the volume of the regional brain subfield of interest, with separate models fitted iteratively for each subregion. *Volume:medication* and *Volume:time* interaction terms were included to test whether regional volumes modulate the associations of medication and treatment time with symptoms. All non-binary variables were *z*-scored and statistical significance was assessed after correction for multiple comparisons using FDR, with corrected p-values < 0.05 considered significant.

### Spatial co-location of subregional associations in SSD with neurotransmitter maps

The regional effect-size maps (t-maps) derived from the previous models of diagnosis, age, medication, and symptoms were spatially correlated with normative neurotransmitter receptor and transporter maps. For each neurotransmitter map, 10,000 surrogate maps preserving spatial autocorrelation were generated using the Eigenstrapping [59] procedure (https://github.com/SNG-newy/eigenstrapping), forming a null distribution. Spearman rank correlations between t-maps and receptor maps were evaluated against this null distribution, with p-values defined as the proportion of surrogate correlations exceeding the observed correlation. Finally, p-values < 0.05 were considered significant after FDR correction across neurotransmitter maps.

## RESULTS

### Assessing attrition effects

Potential attrition bias was assessed by comparing baseline clinical characteristics between participants who remained in the study at the 5-year follow-up (n_retained_=152) and those who dropped out (n_dropout_= 181). No significant differences were observed in baseline positive symptom severity (SAPS; T value = 1.32, p = 0.191), negative symptom severity (SANS; T value = −0.62, p = 0.533), or chlorpromazine-equivalent antipsychotic dose (T value = 0.88, p = 0.377).

### Effects of SSD diagnosis and treatment on regional volumes

Compared to HC, patients exhibited widespread bilateral volume reductions across most of the structures examined at baseline (Figure 2, Baseline). Within the thalamus, the largest diagnostic effects were observed in the medial magnocellular subdivision of the mediodorsal nucleus, whereas the lateral pulvinar was the only thalamic region showing a positive association with diagnosis. However, the remaining pulvinar nuclei showed a negative association with the diagnosis. In the hippocampus, diagnostic effects were also extensively distributed throughout the structure, with the largest effects localized in the CA_4_ subfield. Within the amygdala, the largest effects were observed in the accessory basal nucleus. The brainstem also showed widespread volumetric reductions across its subdivisions, whereas no significant effects were detected in the midbrain. Most regions affected by the disease also showed a negative diagnosis:age_inclusion_ interaction, revealing that the reduction associated with SSD becomes more pronounced at older ages.

**Figure 2.**
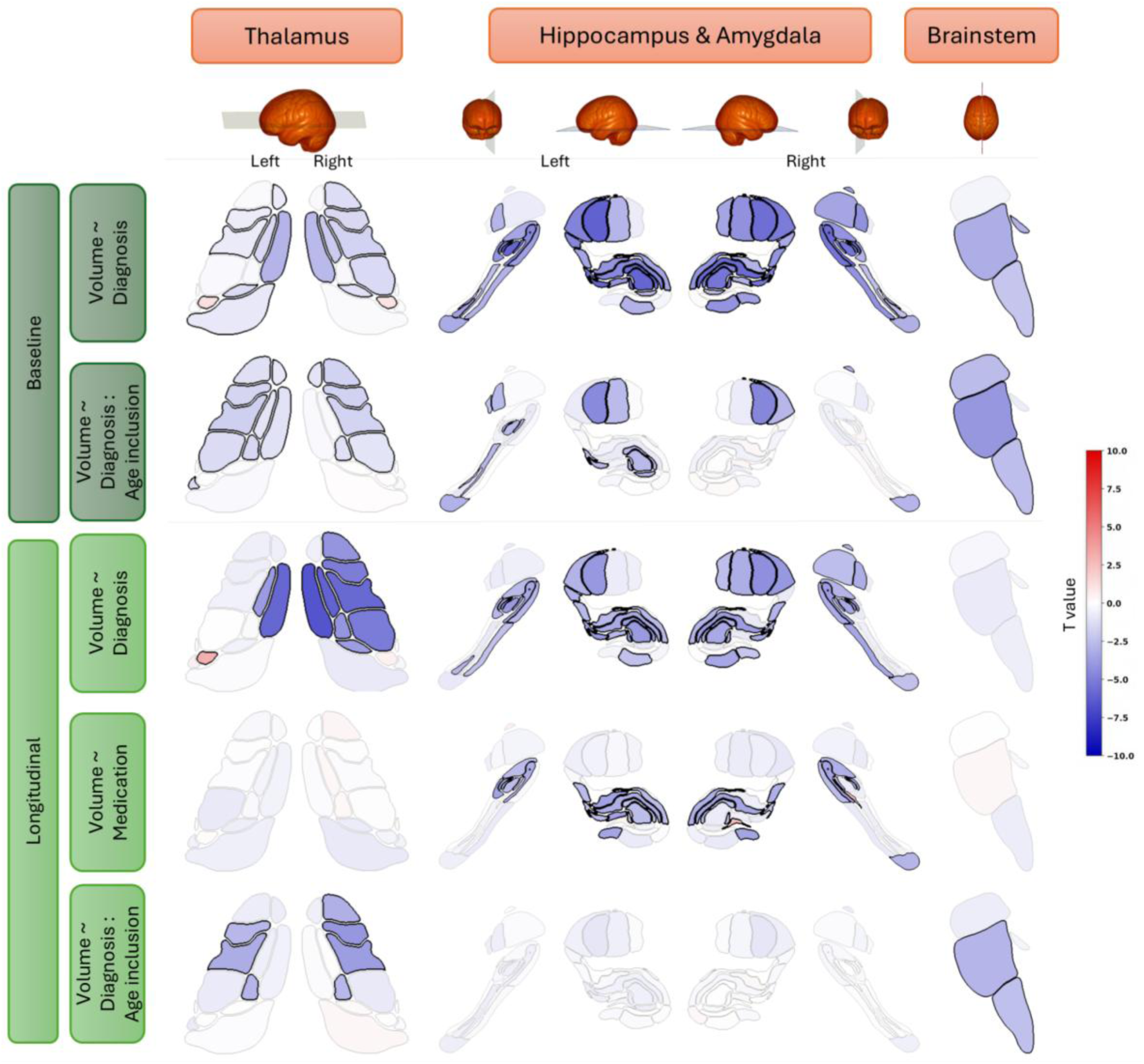
Associations of diagnosis and medication with regional volumes. Contribution of diagnosis, medication and their interaction with age_inclusion_ (rows) to linear models for thalamic, hippocampal, amygdalar, and brainstem subfield volumes (columns). The colour scale ranges from blue (negative T-values, indicating inverse associations with volume) to red (positive T-values, indicating direct associations with volume). Regions that did not survive FDR correction are shown in semi-transparent colours, whereas significant regions are displayed in solid colours. Brain renderings at the top show the anatomical orientation/view for each set of subregions.

The spatial distribution of diagnostic effects in the longitudinal analyses was largely consistent with that observed at baseline, with the exception of the brainstem. Antipsychotic medication was negatively associated with hippocampal subfields but not with thalamus, amygdala, or brainstem (Figure 2, Longitudinal). Within the hippocampus, ventral regions showed the most pronounced deterioration. The diagnosis:age_inclusion_ interaction showed a similar pattern of t-values to that observed in baseline analysis, however, the hippocampal effects did not reach statistical significance. See Supplementary Data for a complete summary of the effects of all variables included across models.

### Regional and subregional volumes are associated with symptomatology

Positive symptoms were strongly and negatively associated with the amygdala volume, with significant associations observed across all subfields except with the Anterior Amygdaloid Area and the Central nucleus (Figure 3). The bilateral Hippocampal-Amygdala Transition Area also showed a strong negative correlation with positive symptomatology (all t-values < −4.8). Several hippocampal subregions were negatively associated with positive symptom severity, with the strongest effects observed in the left body and head of the presubiculum and the parasubiculum (all t-values < −6.5). Interaction effects were widespread, with a positive volume:medication interaction and a negative volume:time interaction across all examined structures. These findings indicate that the negative association between volume and symptoms becomes stronger over the course of treatment but is attenuated in individuals receiving higher doses of antipsychotic medication.

**Figure 3.**
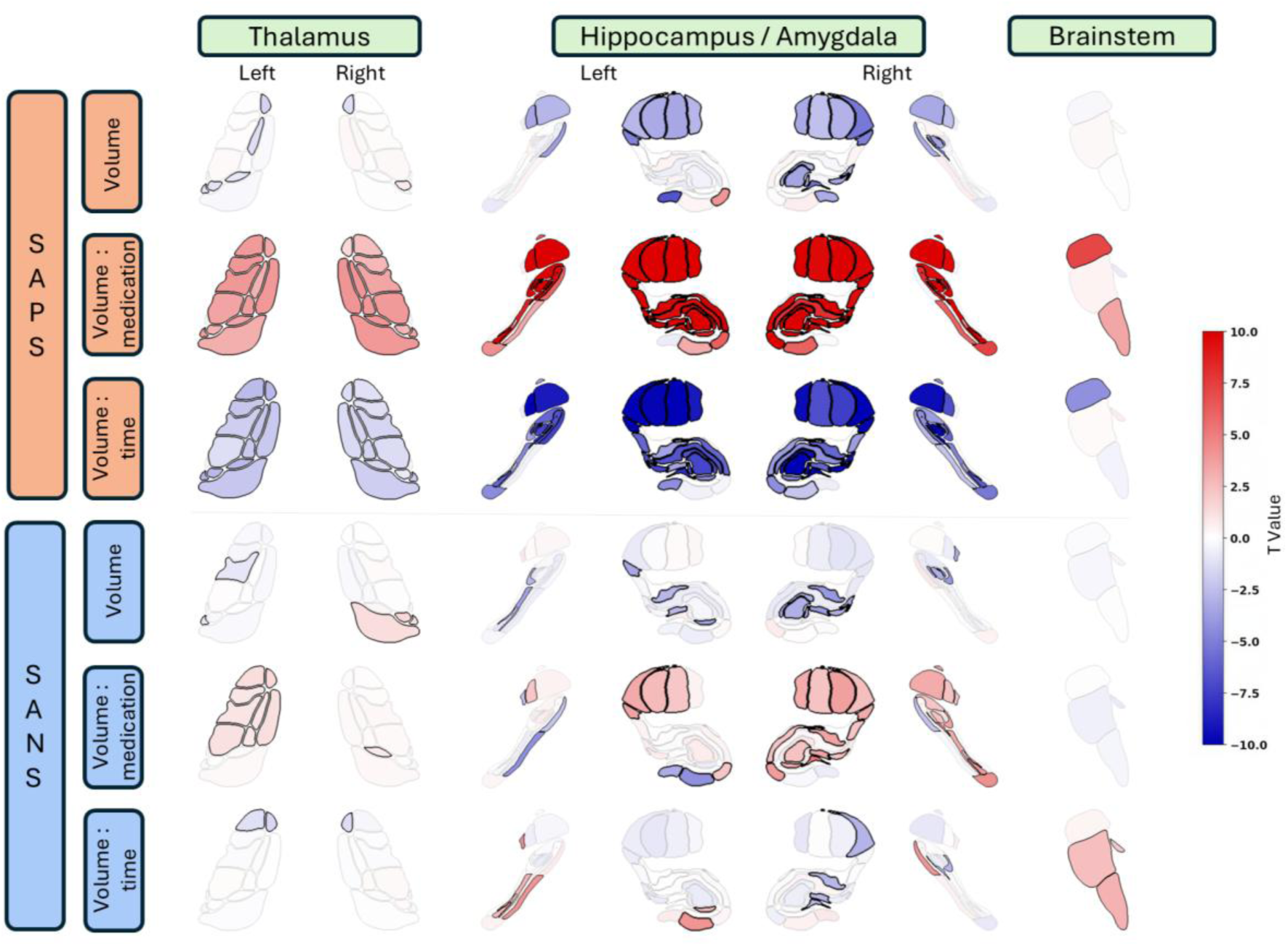
Associations between regional volumes and symptoms. Contributions of regional volume and its interactions with medication and time (rows) to SAPS and SANS associations in linear models for thalamic, hippocampal, amygdalar, and brainstem subfield volumes (columns). The colour scale ranges from blue (negative T-values, indicating inverse associations with symptoms) to red (positive T-values, indicating direct associations with symptoms). Regions that did not survive FDR correction are shown in semi-transparent colours, whereas significant regions are displayed in solid colours.

**Figure 4.**
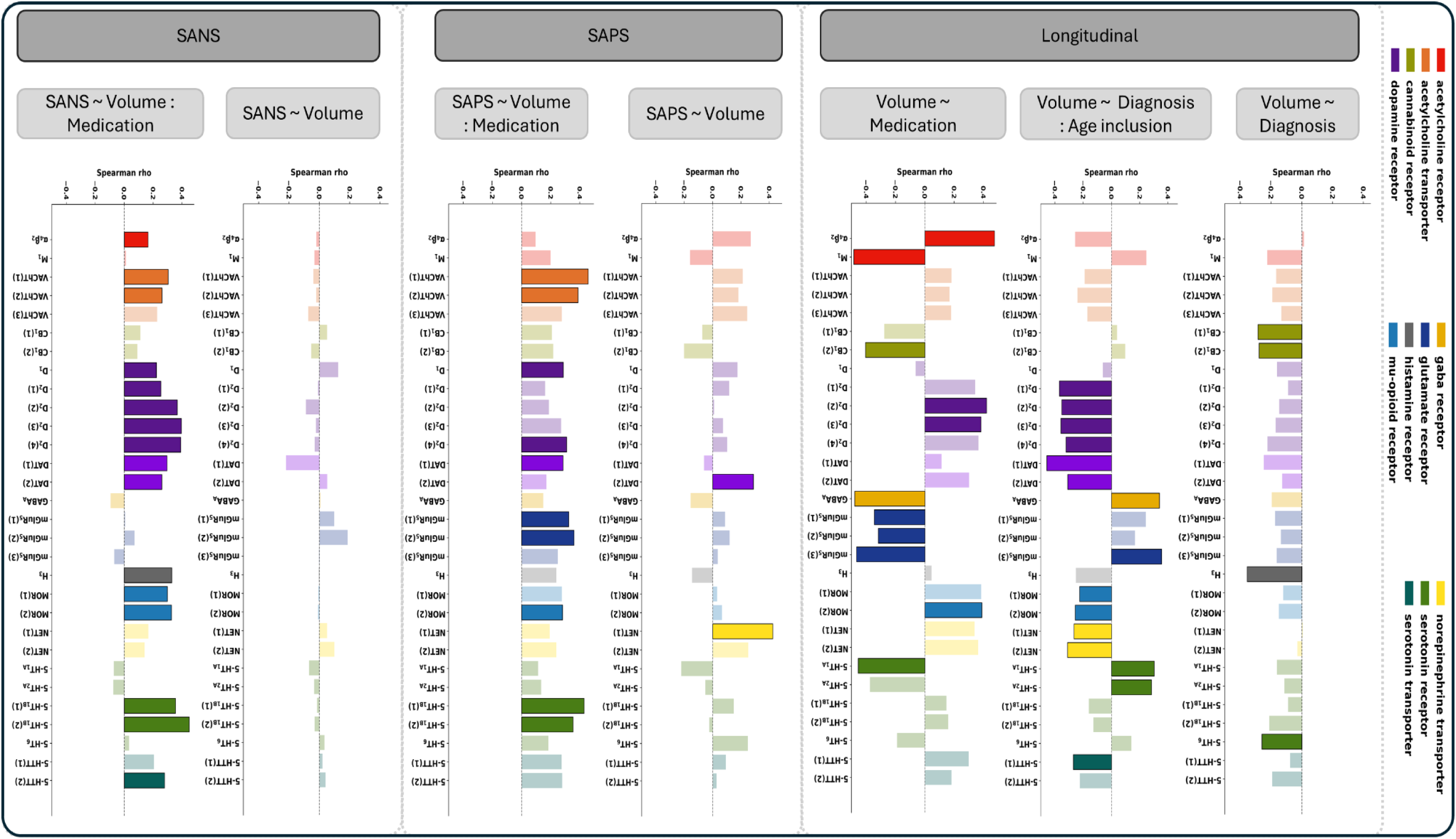
Spatial co-localisation between regional maps of interest derived from the regression models and neurotransmitter distribution maps obtained from normative individuals. Within each panel, each bar represents the Spearman correlation coefficient (ρ) between the regional t-value map corresponding to the indicated model and a neurotransmitter distribution map. Bar colours indicate the target molecule represented by each neurotransmitter map. Bars are shown as solid when the corrected P value is < 0.05 and semi-transparent otherwise.

Negative symptom severity was only modestly associated with studied subcortical volumes, showing a few negative correlations with hippocampal subfields and positive associations with pulvinar regions of the thalamus. The volume:medication interactions were positive but weaker than those observed for positive symptoms, suggesting that medication had a more limited moderating influence on the relationship between volume and negative symptoms. The volume:time interaction also showed weak effects, indicating that treatment time may not modulate the associations between volume and negative symptomatology.

### Spatial co-location of regional associations maps with neurotransmitter distributions

Most t-statistic maps of interest derived from the previous longitudinal models showed significant spatial co-localization with normative neurotransmitter receptor density maps. The map representing the main effect of diagnosis was negatively correlated with CB1, H3, and 5-HT6 receptor density maps (Figure 3A), indicating that the subregions most affected by the disease are those with higher normative densities of cannabinoid, histamine, and serotonin receptors in healthy participants. The diagnosis:age interaction map was negatively co-localized with DAT, D2, H3, MOR, NET, and 5-HTT density maps, suggesting that regions showing age-dependent volume reductions in patients are enriched in dopamine, histamine, cannabinoid, norepinephrine, and serotonin receptors and transporters. The medication-related effect map, with significant negative effects primarily confined to the hippocampus, was negatively co-localized with M1, CB1, GABA, glutamate, and 5-HT1A receptor density maps, indicating that medication-sensitive regions are characterized by higher normative densities of these neurotransmitter systems. In contrast, α4β2, D2, and MOR density maps showed a positive co-localization, suggesting that these medication-sensitive regions have lower normative densities of nicotinic acetylcholine, dopamine D2, and μ-opioid receptors.

Subregional volumes associated with positive symptoms co-localised only with DAT and NET density maps (Figure 3B). In contrast, the volume:medication interaction map showed significant positive co-localisation with VAChT, D1, D2, DAT, mGluR5, MOR, and 5-HT1B density maps. No significant spatial correlations were observed between subregional brain volume and negative symptoms. However, consistent with the findings for positive symptoms, the volume:medication interaction associated with negative symptoms showed widespread positive correlations across a diverse range of neurotransmitter density maps.

Correlation coefficients and empirical p-values for all neurotransmitter and receptor systems are reported in Supplementary Data.

## Discussion

This study characterized cross-sectional and longitudinal volumetric alterations across thalamic, hippocampal, amygdalar, and brainstem subregions in SSD, examined their associations with clinical symptom trajectories, and assessed their spatial correspondence with normative maps of neurotransmitter receptors and transporters.

Our findings indicate that the earliest components of the hippocampal trisynaptic circuit [65], including the molecular layer, dentate gyrus, CA4, and CA3, are preferentially affected at the first episode. As these subfields constitute the hippocampal input pathway [66, 67], this pattern suggests selective vulnerability early in schizophrenia and is consistent with previous reports describing early abnormalities with little additional volumetric decline after the first psychotic episode [60].

A similarly heterogeneous pattern was observed in the amygdala. Widespread volume reductions were already present at the first episode, consistent with previous reports [61, 62]. The largest differences involved the basal nuclei, particularly the accessory basal nucleus, together with the corticoamygdaloid transition area and the hippocampal-amygdaloid transition area. Given that the basolateral amygdalar complex projects broadly to corticolimbic regions, including the hippocampal formation, abnormal GABAergic and glutamatergic signaling within these pathways has been proposed to disrupt excitation–inhibition balance across interconnected neural circuits [15]. Complementary, structural alterations of specific amygdalar nuclei have been related to abnormalities in functional network connectivity in schizophrenia [63]. In contrast, we found no evidence that antipsychotic doses were associated with amygdala volume. Evidence on the effects of antipsychotic medication on amygdalar morphology is inconsistent [64], with studies reporting both medication-related volumetric associations [65], and no significant relationships [66]. The reasons underlying these discrepant findings remain uncertain, although several contributing factors have been proposed, including differences in antipsychotic class, cumulative medication exposure, duration of untreated psychosis, and other clinical and methodological factors [64].

Given the central role of the amygdala in threat detection and fear processing [67], structural alterations within this region may contribute to the emotional and behavioral responses associated with psychotic experiences. Previous studies have linked amygdalar abnormalities to positive symptoms, particularly delusions and violent behavior [68–70]. Consistent with this, the stronger associations observed in our study between amygdalar volumes and positive symptoms, compared with the weaker relationships with negative symptoms, may reflect a preferential involvement of amygdala-related circuits in specific dimensions of psychotic symptomatology.

The thalamus occupies a central position in cortico-subcortical communication, integrating information across sensory, cognitive, and limbic systems. Evidence suggests that thalamic reductions in schizophrenia may preferentially involve specific nuclei, including higher-order association nuclei such as the mediodorsal nucleus (MD) [3, 71]. The MD is extensively connected with the prefrontal cortex [72] and also participates in broader circuits involving limbic regions and the brainstem [73]. This pattern is consistent with our findings, which identified the MD as the most affected thalamic nucleus in SSD [71]. Previous studies have reported that the thalamus may represent an early site of neurodegeneration, from which pathological changes could subsequently spread to structurally connected regions, including the frontal cortex [74].

Regional thalamic volumes have been associated with positive symptoms [75, 76], negative symptoms [77], and cognitive performance [78], supporting the potential clinical relevance of thalamic subregional abnormalities. In our study, the strongest associations were observed with positive symptoms, whereas relationships with negative symptoms were limited. This pattern may indicate that the structural involvement of specific thalamic subregions is more closely related to particular symptom psychotic dimensions although differences in disease stage, clinical characteristics, and methodological approaches may contribute to variability across studies.

Brainstem volumes were already markedly reduced at the first episode [79–81], with no evidence of medication-related effects during follow-up. Moreover, the midbrain showed the strongest volume:time and volume:medication interactions with positive symptom severity. Although the spatial resolution of our analysis precludes attribution of these associations to specific nuclei, the findings are consistent with the central role of midbrain dopaminergic systems in contemporary models of psychosis [8]. In particular, dysregulation of afferent control over midbrain dopamine neurons has been proposed to contribute to abnormal signalling, providing a potential mechanistic link between dysfunction across hippocampal, thalamic, cortical, and striatal circuits and the emergence of psychotic symptoms. However, our findings should not be interpreted as direct evidence of dopaminergic dysfunction, as the observed associations reflect spatial correspondence between structural alterations and normative patterns of dopamine receptor and transporter distribution, rather than direct measures of dopaminergic activity or signalling.

Recent evidence suggests that integrating normative neurotransmitter maps with structural alterations may help identify mechanistically distinct subtypes of schizophrenia [82]. Accordingly, we found that anatomical patterns associated with diagnosis, age, medication, and symptoms aligned with the normative molecular architecture of the brain. Specifically, the regional pattern associated with illness onset showed a negative spatial correspondence with dopaminergic maps and a positive correspondence with GABAergic maps, whereas medication-related alterations showed the opposite pattern. These findings are consistent with proposed models linking hippocampal inhibitory dysfunction to downstream dopaminergic dysregulation [83, 84].

Symptom-related analyses further suggested that distinct clinical dimensions also aligned with partially dissociable molecular distributions. Specifically, regional alterations associated with positive, but not negative, symptoms showed a direct spatial correspondence with dopamine transporter densities, consistent with the well-established role of dopaminergic dysfunction in psychosis [85]. However, the strongest associations emerged when examining how medication exposure interacts with regional volume to influence symptom severity. These volume:medication interactions co-located with dopaminergic and serotonergic (5-HT1B) receptors for both negative and positive symptoms. As core targets of current antipsychotic treatments, both neurotransmitter systems have long been implicated in the pathophysiology of psychotic disorders [86]. Furthermore, these structural alterations spatially converged with VAChT distribution, which plays a key role in resilience against social stress [75], suggesting a contributing role for cholinergic mechanisms. Finally, the volume:medication interaction spatially mapped onto glutamatergic receptor distributions specifically for positive symptoms, consistent with NMDA receptor hypofunction models [76]. Together, these findings support the emerging view that the brain’s intrinsic molecular architecture shapes the spatial distribution of disease-related structural alterations in the subcortex. Moreover, distinct clinical dimensions appear to be associated with partially dissociable, yet interacting, neurochemical systems [58, 87].

### Limitations

Several limitations should be acknowledged. First, because all patients received antipsychotic treatment, medication effects could not be disentangled from illness-related processes, and associations with medication exposure may partly reflect illness severity, treatment history, or other clinical factors rather than medication itself. Second, chlorpromazine-equivalent doses, while widely used, do not account for differences between antipsychotic agents or pharmacological classes. Finally, volumetric estimates of very small substructures are more susceptible to segmentation errors and partial volume effects and should therefore be interpreted with caution.

### Conclusions

Although examining subcortical structures as whole anatomical units can reveal broad patterns of structural alteration, their segmentation into distinct subregions enables the identification of more spatially specific characterisation of volumetric changes. Our findings demonstrate heterogeneous longitudinal volumetric changes across subregions of the thalamus, hippocampus, amygdala, and brainstem in schizophrenia, with these changes showing associations with symptom dimensions and spatial correspondence with the distribution of neurotransmitter receptors and transporters.

## Data and Code availability

All code and non-clinical data used to perform the analyses can be found at https://github.com/NeuroimagingBrainNetworks/SSD-Subcortical-neurotransmitter.

## Authors contributions

C.A.M. performed data curation, methodological design, data analysis, and drafted the manuscript; N.G.S.M, R.A.B, P.S, C.G, A.P, P.S.Q, L.D, M.M.C, R.A.A, J.V.B, J.S., M.R.V, B.C.F and R.R.G contributed to data acquisition, provided advice on data analysis, and participated in writing and editing the manuscript. R.R.G. also contributed to conceptualization and supervision of the work. All authors approved the submitted version of the manuscript.

## References

1. Howes OD, Bukala BR, Beck K. Schizophrenia: from neurochemistry to circuits, symptoms and treatments. Nat Rev Neurol. 2024;20:22–35.

2. Pergola G, Selvaggi P, Trizio S, Bertolino A, Blasi G. The role of the thalamus in schizophrenia from a neuroimaging perspective. Neuroscience & Biobehavioral Reviews. 2015;54:57–75.

3. Zeng V, Hoang D, Song SH, Trotti R, Parker D, Raymond N, et al. Mediodorsal and pulvinar thalamus reductions across the psychosis spectrum. Schizophr Res. 2026;291:88–98.

4. Anticevic A, Halassa MM. The thalamus in psychosis spectrum disorder. Front Neurosci. 2023;17:1163600.

5. Ocak M, Oguz B. Measurement of Limbic System Anatomical Volumes in Patients Diagnosed with Schizophrenia Using Vol2brain and Comparison with Healthy Individuals. Medicina (Kaunas). 2025;61:525.

6. Ohi K, Ishibashi M, Torii K, Hashimoto M, Yano Y, Shioiri T. Differences in subcortical brain volumes among patients with schizophrenia and bipolar disorder and healthy controls. J Psychiatry Neurosci. 2022;47:E77–E85.

7. Tuppurainen H, Kuikka JT, Laakso MP, Viinamäki H, Husso M, Tiihonen J. Midbrain dopamine D2/3 receptor binding in schizophrenia. Eur Arch Psychiatry Clin Neurosci. 2006;256:382–387.

8. Sonnenschein SF, Gomes FV, Grace AA. Dysregulation of Midbrain Dopamine System and the Pathophysiology of Schizophrenia. Front Psychiatry. 2020;11:613.

9. Nopoulos PC, Ceilley JW, Gailis EA, Andreasen NC. An MRI study of midbrain morphology in patients with schizophrenia: relationship to psychosis, neuroleptics, and cerebellar neural circuitry. Biol Psychiatry. 2001;49:13–19.

10. Alsema AM, Puvogel S, Kracht L, Webster MJ, Shannon Weickert C, Eggen BJL, et al. Schizophrenia-associated changes in neuronal subpopulations in the human midbrain. Brain. 2025;148:1374–1388.

11. Kubota M, Miyata J, Sasamoto A, Sugihara G, Yoshida H, Kawada R, et al. Thalamocortical disconnection in the orbitofrontal region associated with cortical thinning in schizophrenia. JAMA Psychiatry. 2013;70:12–21.

12. Hamoda HM, Makhlouf AT, Fitzsimmons J, Rathi Y, Makris N, Mesholam-Gately RI, et al. Abnormalities in thalamo-cortical connections in patients with first-episode schizophrenia: a two-tensor tractography study. Brain Imaging Behav. 2019;13:472–481.

13. Kim M, Kim T, Ha M, Oh H, Moon S-Y, Kwon JS. Large-Scale Thalamocortical Triple Network Dysconnectivities in Patients With First-Episode Psychosis and Individuals at Risk for Psychosis. Schizophr Bull. 2023;49:375–384.

14. Zhu T, Wang Z, Zhou C, Fang X, Huang C, Xie C, et al. Meta-analysis of structural and functional brain abnormalities in schizophrenia with persistent negative symptoms using activation likelihood estimation. Front Psychiatry. 2022;13:957685.

15. Benes FM. Amygdalocortical Circuitry in Schizophrenia: From Circuits to Molecules. Neuropsychopharmacol. 2010;35:239–257.

16. Knight S, McCutcheon R, Dwir D, Grace AA, O’Daly O, McGuire P, et al. Hippocampal circuit dysfunction in psychosis. Transl Psychiatry. 2022;12:344.

17. Ji JL, Diehl C, Schleifer C, Tamminga CA, Keshavan MS, Sweeney JA, et al. Schizophrenia Exhibits Bi-directional Brain-Wide Alterations in Cortico-Striato-Cerebellar Circuits. Cereb Cortex. 2019;29:4463–4487.

18. Huang AS, Rogers BP, Woodward ND. Disrupted modulation of thalamus activation and thalamocortical connectivity during dual task performance in schizophrenia. Schizophr Res. 2019;210:270–277.

19. Yao B, Neggers SFW, Kahn RS, Thakkar KN. Altered thalamocortical structural connectivity in persons with schizophrenia and healthy siblings. Neuroimage Clin. 2020;28:102370.

20. Ramsay IS, Mueller B, Ma Y, Shen C, Sponheim SR. Thalamocortical connectivity and its relationship with symptoms and cognition across the psychosis continuum. Psychological Medicine. 2023;53:5582–5591.

21. Pergola G, Danet L, Pitel A-L, Carlesimo GA, Segobin S, Pariente J, et al. The Regulatory Role of the Human Mediodorsal Thalamus. Trends Cogn Sci. 2018;22:1011–1025.

22. Byne W, Buchsbaum MS, Mattiace LA, Hazlett EA, Kemether E, Elhakem SL, et al. Postmortem assessment of thalamic nuclear volumes in subjects with schizophrenia. Am J Psychiatry. 2002;159:59–65.

23. Highley JR, Walker MA, Crow TJ, Esiri MM, Harrison PJ. Low medial and lateral right pulvinar volumes in schizophrenia: a postmortem study. Am J Psychiatry. 2003;160:1177–1179.

24. Huang AS, Rogers BP, Sheffield JM, Jalbrzikowski ME, Anticevic A, Blackford JU, et al. Thalamic Nuclei Volumes in Psychotic Disorders and in Youths With Psychosis Spectrum Symptoms. Am J Psychiatry. 2020;177:1159–1167.

25. Takahashi T, Tsugawa S, Nakajima S, Plitman E, Chakravarty MM, Masuda F, et al. Thalamic and striato-pallidal volumes in schizophrenia patients and individuals at risk for psychosis: A multi-atlas segmentation study. Schizophr Res. 2022;243:268–275.

26. Perez-Rando M, Elvira UKA, García-Martí G, Gadea M, Aguilar EJ, Escarti MJ, et al. Alterations in the volume of thalamic nuclei in patients with schizophrenia and persistent auditory hallucinations. Neuroimage Clin. 2022;35:103070.

27. Perez-Rando M, Elvira UKA, García-Martí G, Gadea M, Aguilar EJ, Escarti MJ, et al. Alterations in the volume of thalamic nuclei in patients with schizophrenia and persistent auditory hallucinations. Neuroimage Clin. 2022;35:103070.

28. Roeske MJ, Konradi C, Heckers S, Lewis AS. Hippocampal volume and hippocampal neuron density, number and size in schizophrenia: a systematic review and meta-analysis of postmortem studies. Mol Psychiatry. 2021;26:3524–3535.

29. Kawano M, Sawada K, Shimodera S, Ogawa Y, Kariya S, Lang DJ, et al. Hippocampal subfield volumes in first episode and chronic schizophrenia. PLoS One. 2015;10:e0117785.

30. Haukvik UK, Tamnes CK, Söderman E, Agartz I. Neuroimaging hippocampal subfields in schizophrenia and bipolar disorder: A systematic review and meta-analysis. Journal of Psychiatric Research. 2018;104:217–226.

31. Calvo A, Roddy DW, Coughlan H, Kelleher I, Healy C, Harley M, et al. Reduced hippocampal volume in adolescents with psychotic experiences: A longitudinal population-based study. PLoS One. 2020;15:e0233670.

32. Vargas T, Dean DJ, Osborne KJ, Gupta T, Ristanovic I, Ozturk S, et al. Hippocampal Subregions Across the Psychosis Spectrum. Schizophr Bull. 2018;44:1091–1099.

33. Heckers S, Konradi C. GABAergic mechanisms of hippocampal hyperactivity in schizophrenia. Schizophr Res. 2015;167:4–11.

34. Šimić G, Tkalčić M, Vukić V, Mulc D, Španić E, Šagud M, et al. Understanding Emotions: Origins and Roles of the Amygdala. Biomolecules. 2021;11:823.

35. Ho NF, Li Hui Chong P, Lee DR, Chew QH, Chen G, Sim K. The Amygdala in Schizophrenia and Bipolar Disorder: A Synthesis of Structural MRI, Diffusion Tensor Imaging, and Resting-State Functional Connectivity Findings. Harv Rev Psychiatry. 2019;27:150–164.

36. Hoang D, Lizano P, Lutz O, Zeng V, Raymond N, Miewald J, et al. Thalamic, Amygdalar, and hippocampal nuclei morphology and their trajectories in first episode psychosis: A preliminary longitudinal study✰. Psychiatry Res Neuroimaging. 2021;309:111249.

37. Liang S, Wu Y, Hanxiaoran L, Greenshaw AJ, Li T. Anhedonia in Depression and Schizophrenia: Brain Reward and Aversion Circuits. Neuropsychiatr Dis Treat. 2022;18:1385–1396.

38. Perez-Rando M, Penades-Gomiz C, Martinez-Marin P, García-Martí G, Aguilar EJ, Escarti MJ, et al. Volume alterations of the hippocampus and amygdala in patients with schizophrenia and persistent auditory hallucinations. Spanish Journal of Psychiatry and Mental Health. 2025;18:241–249.

39. Kuang Q, Zhou S, Deng G, Zhou N, Zeng X, Zhang H, et al. Subregional amygdala functional connectivity abnormalities and anhedonia impairments in first-episode schizophrenia. BMC Psychiatry. 2025;25:960.

40. Anticevic A, Haut K, Murray JD, Repovs G, Yang GJ, Diehl C, et al. Association of Thalamic Dysconnectivity and Conversion to Psychosis in Youth and Young Adults at Elevated Clinical Risk. JAMA Psychiatry. 2015;72:882–891.

41. Schmitz TW, Correia MM, Ferreira CS, Prescot AP, Anderson MC. Hippocampal GABA enables inhibitory control over unwanted thoughts. Nat Commun. 2017;8:1311.

42. Noga BR, Opris I, Lebedev MA, Mitchell GS. Editorial: Neuromodulatory Control of Brainstem Function in Health and Disease. Front Neurosci. 2020;14:86.

43. Alemán-Morillo C, García-San-Martín N, Bethlehem RA, Segura P, Gomez C, Pasquini A, et al. Early deviations from normative brain morphology and cortical microstructure in schizophrenia spectrum disorders. 2026:2026.01.09.26343760.

44. Lányi O, Zahemszky D, Wenning AS, Engh MA, Molnár Z, Horváth AA, et al. Cerebello-Thalamo-Cortical Dysconnectivity in Schizophrenia Spectrum Disorders: A Resting-State Functional Magnetic Resonance Imaging Meta-Analysis. Biol Psychiatry Cogn Neurosci Neuroimaging. 2026;11:329–346.

45. Gur RE. Pathways to Psychosis: Factors Associated With Risk for Psychosis Spectrum Disorders. Am J Psychiatry. 2025;182:984–990.

46. Di Francesco A, Cutrufelli P, Chiarenza C, Zambuto L, Concerto C, Mineo L, et al. Cannabidiol for the treatment of positive and negative symptoms in schizophrenia spectrum disorders: a systematic review and meta-analysis. Eur Arch Psychiatry Clin Neurosci. 2026. 28 April 2026. 10.1007/s00406-026-02249-3.

47. Panganiban KJ, Smith ECC, Stogios N, Agarwal SM, Ward K, Hahn MK. The cognitive metabolomic signatures in schizophrenia spectrum disorders: A systematic review. Psychiatry Res. 2025;353:116742.

48. Reuter M, Schmansky NJ, Rosas HD, Fischl B. Within-subject template estimation for unbiased longitudinal image analysis. Neuroimage. 2012;61:1402–1418.

49. Gardner M, Shinohara RT, Bethlehem RAI, Romero-Garcia R, Warrier V, Dorfschmidt L, et al. ComBatLS: A Location- and Scale-Preserving Method for Multi-Site Image Harmonization. Human Brain Mapping. 2025;46:e70197.

50. Iglesias JE, Van Leemput K, Augustinack J, Insausti R, Fischl B, Reuter M, et al. Bayesian longitudinal segmentation of hippocampal substructures in brain MRI using subject-specific atlases. Neuroimage. 2016;141:542–555.

51. Iglesias JE, Insausti R, Lerma-Usabiaga G, Bocchetta M, Van Leemput K, Greve DN, et al. A probabilistic atlas of the human thalamic nuclei combining ex vivo MRI and histology. Neuroimage. 2018;183:314–326.

52. Iglesias JE, Augustinack JC, Nguyen K, Player CM, Player A, Wright M, et al. A computational atlas of the hippocampal formation using ex vivo, ultra-high resolution MRI: Application to adaptive segmentation of in vivo MRI. NeuroImage. 2015;115:117–137.

53. Saygin ZM, Kliemann D, Iglesias JE, van der Kouwe AJW, Boyd E, Reuter M, et al. High-resolution magnetic resonance imaging reveals nuclei of the human amygdala: manual segmentation to automatic atlas. Neuroimage. 2017;155:370–382.

54. Iglesias JE, Van Leemput K, Bhatt P, Casillas C, Dutt S, Schuff N, et al. Bayesian segmentation of brainstem structures in MRI. Neuroimage. 2015;113:184–195.

55. Andreasen NC. The Scale for the Assessment of Negative Symptoms (SANS): conceptual and theoretical foundations. Br J Psychiatry Suppl. 1989:49–58.

56. Andreasen NC. Scale for the assessment of positive symptoms. Group. 1984;17:173–180.

57. Rodriguez-Perez N, Ayesa-Arriola R, Ortiz-García de la Foz V, Setien-Suero E, Tordesillas-Gutierrez D, Crespo-Facorro B. Long term cortical thickness changes after a first episode of non-affective psychosis: The 10 year follow-up of the PAFIP cohort. Progress in Neuro-Psychopharmacology and Biological Psychiatry. 2021;108:110180.

58. Markello RD, Hansen JY, Liu Z-Q, Bazinet V, Shafiei G, Suárez LE, et al. neuromaps: structural and functional interpretation of brain maps. Nat Methods. 2022;19:1472–1479.

59. Koussis NC, Pang JC, Phogat R, Jeganathan J, Paton B, Fornito A, et al. Generation of surrogate brain maps preserving spatial autocorrelation through random rotation of geometric eigenmodes. Imaging Neurosci (Camb). 2025;3:IMAG.a.71.

60. Chopra S, Segal A, Oldham S, Holmes A, Sabaroedin K, Orchard ER, et al. Network-Based Spreading of Gray Matter Changes Across Different Stages of Psychosis. JAMA Psychiatry. 2023;80:1246–1257.

61. Barth C, Nerland S, de Lange A-MG, Wortinger LA, Hilland E, Andreassen OA, et al. In Vivo Amygdala Nuclei Volumes in Schizophrenia and Bipolar Disorders. Schizophr Bull. 2021;47:1431–1441.

62. Zheng F, Li C, Zhang D, Cui D, Wang Z, Qiu J. Study on the sub-regions volume of hippocampus and amygdala in schizophrenia. Quant Imaging Med Surg. 2019;9:1025–1036.

63. Alharthi RR, Banaja D, Alahmadi A, Alsalah JH, Baeshen A, Alghamdi AH, et al. Altered Functional Connectivity of Amygdala Subregions with Large-Scale Brain Networks in Schizophrenia: A Resting-State fMRI Study. Tomography. 2026;12:2.

64. Xia M, Wang Y, Su W, Tang Y, Zhang T, Cui H, et al. The effect of initial antipsychotic treatment on hippocampal and amygdalar volume in first-episode schizophrenia is influenced by age. Progress in Neuro-Psychopharmacology and Biological Psychiatry. 2023;126:110780.

65. Poeppl TB, Frank E, Schecklmann M, Kreuzer PM, Prasser SJ, Rupprecht R, et al. Amygdalohippocampal neuroplastic changes following neuroleptic treatment with quetiapine in first-episode schizophrenia. Int J Neuropsychopharmacol. 2014;17:833–843.

66. Velakoulis D, Wood SJ, Wong MTH, McGorry PD, Yung A, Phillips L, et al. Hippocampal and amygdala volumes according to psychosis stage and diagnosis: a magnetic resonance imaging study of chronic schizophrenia, first-episode psychosis, and ultra-high-risk individuals. Arch Gen Psychiatry. 2006;63:139–149.

67. Kosteletou-Kassotaki E, Cinca-Tomás MT, Varriano F, Soria G, Prats-Galino A, Domínguez-Borràs J. A Direct Auditory Subcortical Route to the Amygdala Associated with Fear in Humans. J Neurosci. 2026;46.

68. Feng X, Provenzano F, Appelbaum PS, Masucci MD, Brucato G, Lieberman JA, et al. Amygdalar Volume and Violent Ideation in a Sample at Clinical High-Risk for Psychosis. Psychiatry Res Neuroimaging. 2019;287:60–62.

69. Bell C, Tesli N, Gurholt TP, Rokicki J, Hjell G, Fischer-Vieler T, et al. Associations between amygdala nuclei volumes, psychosis, psychopathy, and violent offending. Psychiatry Research: Neuroimaging. 2022;319:111416.

70. García-San-Martín N, Bethlehem RA, Sebenius I, Saraiva LC, Segura P, Alemán-Morillo C, et al. Long-term morphometric similarity gradients relate to cortical hierarchy and psychiatric symptoms in schizophrenia. 2026:2026.02.25.26347075.

71. Young KA, Manaye KF, Liang C, Hicks PB, German DC. Reduced number of mediodorsal and anterior thalamic neurons in schizophrenia. Biol Psychiatry. 2000;47:944–953.

72. Parnaudeau S, Bolkan SS, Kellendonk C. The Mediodorsal Thalamus: An Essential Partner of the Prefrontal Cortex for Cognition. Biol Psychiatry. 2018;83:648–656.

73. Marcuse LV, Langan M, Hof PR, Panov F, Saez I, Jimenez-Shahed J, et al. The thalamus: Structure, function, and neurotherapeutics. Neurotherapeutics. 2025;22:e00550.

74. Jiang Y, Luo C, Li X, Duan M, He H, Chen X, et al. Progressive Reduction in Gray Matter in Patients with Schizophrenia Assessed with MR Imaging by Using Causal Network Analysis. Radiology. 2018;287:633–642.

75. Ferri J, Ford JM, Roach BJ, Turner JA, van Erp TG, Voyvodic J, et al. Resting-state thalamic dysconnectivity in schizophrenia and relationships with symptoms. Psychol Med. 2018;48:2492–2499.

76. Rao NP, Kalmady S, Arasappa R, Venkatasubramanian G. Clinical correlates of thalamus volume deficits in anti-psychotic-naïve schizophrenia patients: A 3-Tesla MRI study. Indian J Psychiatry. 2010;52:229–235.

77. Bayrakçı A, Zorlu N, Karakılıç M, Gülyüksel F, Yalınçetin B, Oral E, et al. Negative symptoms are associated with modularity and thalamic connectivity in schizophrenia. Eur Arch Psychiatry Clin Neurosci. 2023;273:565–574.

78. Thalhammer M, Schulz J, Scheulen F, Oubaggi MEM, Kirschner M, Kaiser S, et al. Distinct Volume Alterations of Thalamic Nuclei Across the Schizophrenia Spectrum. Schizophr Bull. 2024;50:1208–1222.

79. Aoyama S, Okuda H, Furuzawa N, Yoneda H, Fujikane D, Takai K, et al. Sex differences in brainstem structure volumes in patients with schizophrenia. Schizophrenia (Heidelb). 2023;9:16.

80. Fritze S, Harneit A, Waddington JL, Kubera KM, Schmitgen MM, Otte M-L, et al. Structural alterations in brainstem, basal ganglia and thalamus associated with parkinsonism in schizophrenia spectrum disorders. Eur Arch Psychiatry Clin Neurosci. 2021;271:1455–1464.

81. Hirjak D, Wolf RC, Stieltjes B, Hauser T, Seidl U, Thiemann U, et al. Neurological soft signs and brainstem morphology in first-episode schizophrenia. Neuropsychobiology. 2013;68:91–99.

82. Hahn L, Raabe FJ, Keeser D, Vetter C, Fanning J, Hasan A, et al. Mapping Neurochemical Signatures onto Brain Structure for Neurotransmitter-Informed Discrimination of Schizophrenia Patients from Healthy Controls. 2025.

83. Lodge DJ, Grace AA. Hippocampal dysregulation of dopamine system function and the pathophysiology of schizophrenia. Trends Pharmacol Sci. 2011;32:507–513.

84. Grace AA. Dysregulation of the dopamine system in the pathophysiology of schizophrenia and depression. Nat Rev Neurosci. 2016;17:524–532.

85. Meyer JM. How antipsychotics work in schizophrenia: a primer on mechanisms. CNS Spectr. 2024;30:e6.

86. Leucht S, Priller J, Davis JM. Antipsychotic Drugs: A Concise Review of History, Classification, Indications, Mechanism, Efficacy, Side Effects, Dosing, and Clinical Application. American Journal of Psychiatry. 2024;181:865–878.

87. Hansen JY, Shafiei G, Markello RD, Smart K, Cox SML, Nørgaard M, et al. Mapping neurotransmitter systems to the structural and functional organization of the human neocortex. Nat Neurosci. 2022;25:1569–1581.

