## Supplemental Material for "Thalamic, hippocampal, amygdalar and brainstem long-term changes in schizophrenia co-localised with normative neurotransmitter distributions"

#### Supplementary Material

### Participants

The study included individuals aged 15 to 60 years presenting with a first episode of psychosis (FEP), who resided within the catchment area and had received either no antipsychotic treatment or less than six weeks of exposure before enrolment. Participants with DSM-IV diagnoses of alcohol or drug dependence, intellectual disability, neurological disorders, or a history of significant head injury were excluded. Clinical diagnoses were established by an experienced psychiatrist using the Structured Clinical Interview for DSM-IV Axis I Disorders (SCID-I) within six months of the first clinical presentation.

Participants were recruited through PAFIP (Programa de Atención a las Fases Iniciales de Psicosis), a multidisciplinary early intervention program based at the University Hospital Marqués de Valdecilla, which serves an epidemiologically representative population in Cantabria, Spain. Referrals were received from psychiatric inpatient units, emergency departments, community mental health services, and other healthcare providers throughout the region.

Healthy controls were recruited from the same geographical area and screened using a shortened version of the Comprehensive Assessment of Symptoms and History to exclude psychiatric and neurological disorders, major medical illness, and substance dependence.

### Supplementary tables

| Diagnosis | Sex | Number | Age at inclusion |
|---|---|---|---|
| Controls | Male | 120 | 28.4 ± 7.06 |
| | Female | 75 | 30.1 ± 8.39 |
| SSD | Male | 213 | 27.6 ± 7.46 |
| | Female | 144 | 33.0 ± 9.55 |

**Table S1.** Demographic characteristics and number of healthy controls and participants with schizophrenia spectrum disorders (SSD), stratified by sex. Age at study inclusion is presented as mean ± standard deviation.

| Symptom | | Male | Female |
|---|---|---|---|
| Positive symptoms | Hallucinations | 2.50 ± 2.33 | 2.22 ± 2.28 |
| | Delusions | 4.58 ± 1.25 | 4.61 ± 1.09 |
| | Bizarre Behaviour | 3.91 ± 1.71 | 4.10 ± 1.43 |
| | Inappropriate Affect | 1.20 ± 1.76 | 1.47 ± 1.92 |
| | Disorganized Thinking | 0.91 ± 1.64 | 1.24 ± 1.79 |
| | SAPS Total Score | 13.10 ± 5.26 | 13.65 ± 5.28 |

| Symptom | | Male | Female |
|---|---|---|---|
| Negative symptoms | Affective Flattening | 1.25 ± 1.50 | 0.83 ± 1.36 |
| | Alogia | 0.83 ± 1.45 | 0.68 ± 1.36 |
| | Anhedonia | 1.56 ± 1.80 | 1.04 ± 1.62 |
| | Attention | 1.71 ± 1.94 | 2.03 ± 1.95 |
| | Avolition | 1.09 ± 1.68 | 0.74 ± 1.43 |
| | SANS Total Score | 6.43 ± 6.05 | 5.07 ± 5.04 |

**Table S2.** Positive and negative symptom profiles at study entry according to sex. Values are expressed as mean ± standard deviation. SAPS = Scale for the Assessment of Positive Symptoms; SANS = Scale for the Assessment of Negative Symptoms.

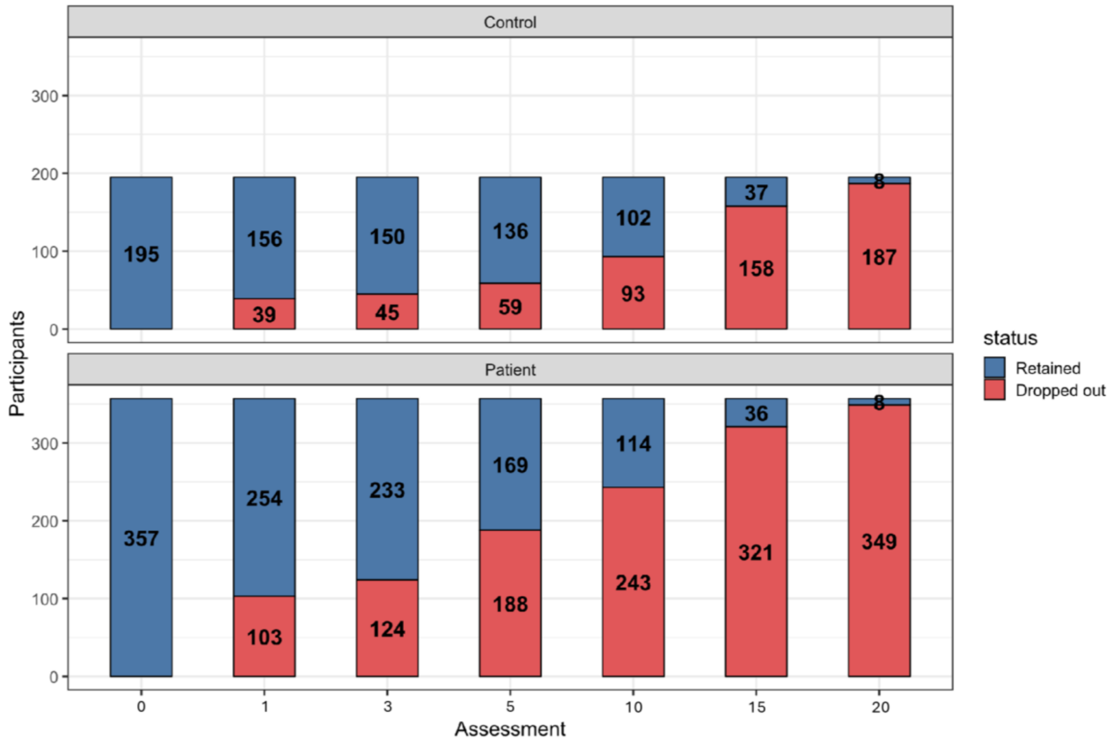

**Table S3.** Retained and dropped out participant number across the study.

| Neurotransmitter | Molecular Target | Reference |
|---|---|---|
| Acetylcholine | α4β2 receptor | Hillmer et al., 2016 [1] |
| Acetylcholine | M1 receptor | Naganawa et al., 2020 [2] |
| Acetylcholine | VAChT | Aghourian et al., 2017 [3] |
| Acetylcholine | VAChT | Bedard et al., 2019 [4] |
| Acetylcholine | VAChT | Tuominen et al. [5] |
| Cannabinoid | CB1 receptor | Laurikainen et al., 2018 [6] |
| Cannabinoid | CB1 receptor | et al., 2015 [7] |
| Dopamine | D1 receptor | Kaller et al., 2017 [8] |
| Dopamine | D2 receptor | Alakurtti et al., 2015 [9] |
| Dopamine | D2 receptor | Jaworska et al., 2020 [10] |
| Dopamine | D2 receptor | Sandiego et al., 2015 [11] |
| Dopamine | D2 receptor | Smith et al., 2017 [12] |
| Dopamine | DAT | Dukart et al., 2018 [13] |
| Dopamine | DAT | Sasaki et al., 2012 [14] |
| GABA | GABA(_A) receptor | Dukart et al., 2018 [13] |
| Glutamate | mGluR5 | Dubois et al., 2015 [15] |
| Glutamate | mGluR5 | Rosaneto et al. [16] |
| Glutamate | mGluR5 | Smart et al., 2019 [17] |
| Histamine | H3 receptor | Gallezot et al., 2017 [18] |
| Opioid | μ-opioid receptor (MOR) | Kantonen et al., 2020 [19] |
| Opioid | μ-opioid receptor (MOR) | Turtonen et al., 2020 [20] |
| Noradrenaline | NET | Ding et al., 2010 [21] |
| Noradrenaline | NET | Hesse et al., 2017 [22] |
| Serotonin | 5-HT1A receptor | Savli et al., 2012 [23] |
| Serotonin | 5-HT2A receptor | Savli et al., 2012 [23] |
| Serotonin | 5-HT1B receptor | Savli et al., 2012 [23] |
| Serotonin | 5-HT1B receptor | Gallezot et al., 2010 [18] |
| Serotonin | 5-HT6 receptor | Radhakrishnan et al., 2018 [24] |
| Serotonin | 5-HTT | Fazio et al., 2016 [25] |
| Serotonin | 5-HTT | Savli et al., 2012 [23] |

**Table S4.** Neuromaps-derived brain maps included in the analyses. For each map, the corresponding neurotransmitter system (spatial distribution), the target molecule used for map generation, and the original reference from which the map was obtained are reported.

| Variable | Retained (n = 152) | Dropped out (n = 181) | p-value | T value |
|---|---|---|---|---|
| SAPS | 13.37 ± 4.25 | 14.13 ± 4.54 | 0.191 | 1.32 |
| SANS | 6.28 ± 5.06 | 5.84 ± 5.93 | 0.533 | -0.62 |
| Chlorpromazine equivalents | 188.08 ± 95.65 | 199.00 ± 77.06 | 0.377 | 0.88 |

**Table S5.** Comparison of baseline characteristics between retained and dropout patients (at last 5 years in the study).

### References

1. Hillmer AT, Esterlis I, Gallezot JD, Bois F, Zheng MQ, Nabulsi N, et al. Imaging of cerebral α4β2* nicotinic acetylcholine receptors with (-)-[(18)F]Flubatine PET: Implementation of bolus plus constant infusion and sensitivity to acetylcholine in human brain. Neuroimage. 2016;141:71–80.
2. Naganawa M, Nabulsi N, Henry S, Matuskey D, Lin S-F, Slieker L, et al. First-in-Human Assessment of 11C-LSN3172176, an M1 Muscarinic Acetylcholine Receptor PET Radiotracer. J Nucl Med. 2021;62:553–560.
3. Aghourian M, Legault-Denis C, Soucy J-P, Rosa-Neto P, Gauthier S, Kostikov A, et al. Quantification of brain cholinergic denervation in Alzheimer's disease using PET imaging with [18F]-FEOBV. Mol Psychiatry. 2017;22:1531–1538.
4. Bedard M-A, Aghourian M, Legault-Denis C, Postuma RB, Soucy J-P, Gagnon J-F, et al. Brain cholinergic alterations in idiopathic REM sleep behaviour disorder: a PET imaging study with 18F-FEOBV. Sleep Med. 2019;58:35–41.
5. Hansen JY, Shafiei G, Markello RD, Smart K, Cox SML, Nørgaard M, et al. Mapping neurotransmitter systems to the structural and functional organization of the human neocortex. Nat Neurosci. 2022;25:1569–1581.
6. Laurikainen H, Tuominen L, Tikka M, Merisaari H, Armio R-L, Sormunen E, et al. Sex difference in brain CB1 receptor availability in man. Neuroimage. 2019;184:834–842.
7. Normandin MD, Zheng M-Q, Lin K-S, Mason NS, Lin S-F, Ropchan J, et al. Imaging the cannabinoid CB1 receptor in humans with [11C]OMAR: assessment of kinetic analysis methods, test-retest reproducibility, and gender differences. J Cereb Blood Flow Metab. 2015;35:1313–1322.
8. Kaller S, Rullmann M, Patt M, Becker G-A, Luthardt J, Girbardt J, et al. Test–retest measurements of dopamine D1-type receptors using simultaneous PET/MRI imaging. Eur J Nucl Med Mol Imaging. 2017;44:1025–1032.
9. Alakurtti K, Johansson JJ, Joutsa J, Laine M, Bäckman L, Nyberg L, et al. Long-term test–retest reliability of striatal and extrastriatal dopamine D2/3 receptor binding: study with [11C]raclopride and high-resolution PET. J Cereb Blood Flow Metab. 2015;35:1199–1205.
10. Jaworska N, Cox SML, Tippler M, Castellanos-Ryan N, Benkelfat C, Parent S, et al. Extra-striatal D2/3 receptor availability in youth at risk for addiction. Neuropsychopharmacology. 2020;45:1498–1505.
11. Sandiego CM, Gallezot J-D, Lim K, Ropchan J, Lin S, Gao H, et al. Reference region modeling approaches for amphetamine challenge studies with [11C]FLB 457 and PET. J Cereb Blood Flow Metab. 2015;35:623–629.
12. Smith CT, Crawford JL, Dang LC, Seaman KL, San Juan MD, Vijay A, et al. Partial-volume correction increases estimated dopamine D2-like receptor binding potential and reduces adult age differences. J Cereb Blood Flow Metab. 2019;39:822–833.
13. Dukart J, Holiga Š, Chatham C, Hawkins P, Forsyth A, McMillan R, et al. Cerebral blood flow predicts differential neurotransmitter activity. Sci Rep. 2018;8:4074.
14. Sasaki T, Ito H, Kimura Y, Arakawa R, Takano H, Seki C, et al. Quantification of dopamine transporter in human brain using PET with 18F-FE-PE2I. J Nucl Med. 2012;53:1065–1073.
15. DuBois JM, Rousset OG, Rowley J, Porras-Betancourt M, Reader AJ, Labbe A, et al. Characterization of age/sex and the regional distribution of mGluR5

availability in the healthy human brain measured by high-resolution [(11)C]ABP688 PET. Eur J Nucl Med Mol Imaging. 2016;43:152–162.
16. Hansen JY, Shafiei G, Markello RD, Smart K, Cox SML, Nørgaard M, et al. Mapping neurotransmitter systems to the structural and functional organization of the human neocortex. Nat Neurosci. 2022;25:1569–1581.
17. Smart K, Cox SML, Scala SG, Tippler M, Jaworska N, Boivin M, et al. Sex differences in [11C]ABP688 binding: a positron emission tomography study of mGlu5 receptors. Eur J Nucl Med Mol Imaging. 2019;46:1179–1183.
18. Gallezot J-D, Nabulsi N, Neumeister A, Planeta-Wilson B, Williams WA, Singhal T, et al. Kinetic modeling of the serotonin 5-HT1B receptor radioligand [11C]P943 in humans. J Cereb Blood Flow Metab. 2010;30:196–210.
19. Kantonen T, Karjalainen T, Isojärvi J, Nuutila P, Tuisku J, Rinne J, et al. Interindividual variability and lateralization of μ-opioid receptors in the human brain. Neuroimage. 2020;217:116922.
20. Turtonen O, Saarinen A, Nummenmaa L, Tuominen L, Tikka M, Armio R-L, et al. Adult Attachment System Links With Brain Mu Opioid Receptor Availability In Vivo. BPS: CNNI. 2021;6:360–369.
21. Ding Y-S, Singhal T, Planeta-Wilson B, Gallezot J-D, Nabulsi N, Labaree D, et al. PET imaging of the effects of age and cocaine on the norepinephrine transporter in the human brain using (S,S)-[(11)C]O-methylreboxetine and HRRT. Synapse. 2010;64:30–38.
22. Hesse S, Becker G-A, Rullmann M, Bresch A, Luthardt J, Hankir MK, et al. Central noradrenaline transporter availability in highly obese, non-depressed individuals. Eur J Nucl Med Mol Imaging. 2017;44:1056–1064.
23. Savli M, Bauer A, Mitterhauser M, Ding Y-S, Hahn A, Kroll T, et al. Normative database of the serotonergic system in healthy subjects using multi-tracer PET. Neuroimage. 2012;63:447–459.
24. Radhakrishnan R, Nabulsi N, Gaiser E, Gallezot J-D, Henry S, Planeta B, et al. Age-Related Change in 5-HT6 Receptor Availability in Healthy Male Volunteers Measured with 11C-GSK215083 PET. J Nucl Med. 2018;59:1445–1450.
25. Fazio P, Schain M, Varnäs K, Halldin C, Farde L, Varrone A. Mapping the distribution of serotonin transporter in the human brainstem with high-resolution PET: Validation using postmortem autoradiography data. Neuroimage. 2016;133:313–320.
